# Analysis of Errors in Issuer-Side QHP APIs and ACA Marketplace Machine-Readable Files (MRFs) in the United States

**DOI:** 10.64898/2025.12.05.25341700

**Authors:** Dmytro Nikolayev

## Abstract

The Centers for Medicare and Medicaid Services (CMS) requires Qualified Health Plan (QHP) issuers on the federal health insurance marketplace to publish machine-readable JSON files describing plans, provider networks, and drug formularies, following the QHP Provider & Formulary API specification (`index.json`, `plans.json`, `providers.json`, and `drugs.json`). These data are intended to support consumer-facing tools that help people compare coverage options.

In parallel, CMS’s Center for Consumer Information & Insurance Oversight (CCIIO) publishes Health Insurance Exchange Public Use Files (Exchange PUFs) for plan years 2014–2026, including a Machine-readable URL PUF (MR-PUF). The MR-PUF provides issuer-level records with state, a five-digit HIOS issuer ID, the issuer’s machine-readable index URL for provider/formulary/plan JSON, and a technical point-of-contact email, and is available for plan years 2016–2026. For plan year 2026, CMS distributes the MR-PUF as `machine-readable-url-puf.zip` under the 2026 Exchange PUFs.

In this study, we analyze 59,899 import errors recorded by the HealthPorta Healthcare Data Dashboard across 735 issuer import logs, covering QHP provider and formulary APIs as of December 2025. Our analysis focuses on plan years 2025 and 2026, which account for the overwhelming majority of observed errors, but we also document that some issuers still publish data for older plan years (2018–2024) inside the same JSON files. Among the 735 issuers, 339 exhibit at least one error.

We classify error messages into a small set of recurring categories: missing recommended `cost_sharing` fields (48.0% of all errors), missing required `formulary` fields (26.5%), and issuer identifier mismatches relative to the CMS machine-readable URL index (24.8%). A further 0.6% of errors reflect invalid JSON responses, and small residual fractions arise from network/TLS failures, HTTP errors, schema anomalies (e.g., overly long plan IDs), and drug-related field omissions. JSON-related problems include non-UTF-8 bytes, HTML or other non-JSON payloads returned at QHP API endpoints, truncated or incomplete responses, and lexical parse errors.

By aggregating errors by source (`plans.json`, `providers.json`, `drugs.json`, and index files), plan year (with special attention to 2025–2026), issuer, state, and endpoint URL, we show that a narrow set of issuer-side problems accounts for the vast majority of observed failures. We then analyze, using the QHP GitHub specification, the CMS Exchange PUF documentation and the Healthcare MRF API import model, why the missing fields we observe (`formulary`, `cost_sharing`, IDs, tiers, and policy flags) are structurally necessary and how their absence cascades into broken joins and misleading analytics when QHP data are combined with machine-readable file (MRF) rate data. We conclude with concrete recommendations for issuer and vendor pipelines.

## 1 Introduction

Beginning in 2015, CMS and health insurance carriers jointly defined a JSON-based schema for sharing machine-readable information about health care providers and drug formularies covered by Qualified Health Plans (QHPs) on the federal marketplace, building on regulatory standards in 45 CFR 156 (including network adequacy and formulary transparency requirements) [1]. QHP issuers host a set of public JSON documents—commonly `index.json`, `plans.json`, `providers.json`, and `drugs.json`—that describe their marketplace plans, associated provider networks, and drug coverage. These URLs are collected by CMS and made available so that third parties can aggregate the data and build tools that help consumers compare coverage options. Prior empirical work has documented substantial and persistent inaccuracies in health plan provider directories and has linked directory errors to out-of-network care and surprise billing, underscoring the importance of robust machine-readable directory infrastructure [2, 3, 4].

Alongside the APIs themselves, CMS’s Center for Consumer Information & Insurance Oversight (CCIIO) publishes Health Insurance Exchange Public Use Files (Exchange PUFs). These downloadable datasets, available for plan years 2014 through 2026, provide plan- and issuer-level information on certified QHPs and stand-alone dental plans (SADPs) and are explicitly intended to support timely benefit and rate analysis. CMS notes that the 2026 Exchange PUFs are updated so they reflect the plan data consumers see when shopping for QHPs on the Exchanges, with data for the 2026 PUFs imported into CMS systems by late November 2025 [5].

One of the Exchange PUF datasets is the Machine-readable URL PUF (MR-PUF), which contains issuer-level records identifying machine-readable index URLs that lead to each issuer’s provider, formulary, and plan JSON files. The PY26 data dictionary describes four variables for this file: State (two-character state abbreviation), Issuer ID (five-digit HIOS issuer code), URL Submitted (the URL that contains the issuer’s machine-readable provider, formulary, and plan JSON URLs), and Tech POC Email (email of the technical point of contact for the issuer) [6]. CMS indicates that the MR-PUF is available for plan years 2016 through 2026 [5]. For 2026, CMS publishes the MR-PUF as a compressed file at:

https://download.cms.gov/marketplace-puf/2026/machine-readable-url-puf.zip[7].

In parallel, transparency regulations and CMS programs have led to the publication of large machine-readable files (MRFs) describing allowed amounts and negotiated rates. The open-source Healthcare MRF API is one implementation of a full-stack importer and API for marketplace MRFs: it downloads issuer-hosted transparency files, normalizes them into a PostgreSQL database, and exposes the result via REST endpoints [8]. In production, HealthPorta uses this MRF pipeline alongside QHP provider/formulary data and CMS Exchange PUFs to power its ACA Healthcare Data Dashboard and plan-profile pages.

Recent federal health plan price transparency initiatives, including the Transparency in Coverage (TiC) final rule, require most group health plans and issuers of individual coverage to publish machine-readable files with in-network negotiated rates and out-of-network allowed amounts for covered items and services [9]. CMS explicitly anticipates that these files will be consumed primarily by “persons and entities with the requisite experience and expertise”—such as researchers and application developers—who can transform them into tools that make pricing information accessible and useful for individual consumers and other purchasers [10]. In this policy vision, open MRF data are supposed to make the financial side of health care more legible by enabling plan-to-plan comparisons of expected costs, rather than leaving prices buried in unstructured documents.

The QHP APIs, the Exchange PUFs (especially the MR-PUF), and the MRF API interact through shared identifiers and shared semantics: plan IDs, issuer IDs, network tiers, drug tiers, and cost-sharing rules. When QHP fields are missing or inconsistent, they not only break QHP-only tools, but also limit what can be done with PUF- and MRF-derived data that depends on them.

In this paper, we use HealthPorta’s Healthcare Data Dashboard as a large natural experiment in issuer-side data quality for the QHP Provider & Formulary APIs. The dashboard performs nightly imports of QHP API JSON files and records detailed error messages when ingestion fails. By aggregating these logs across all issuers tracked by the dashboard, we obtain a cross-sectional view of the operational problems that arise when real-world QHP APIs are ingested in bulk.

Our analysis primarily targets plan years 2025 and 2026, because these correspond to the most recent coverage years at the time of our snapshot and account for nearly all observed errors. Nonetheless, we also find that some issuers continue to publish older plan years (2018–2024) in the same JSON files, leading to a long tail of additional errors for historical data.

Our contributions are threefold:

1. We compile and describe a structured dataset of 59,899 import errors across 735 issuers, with one entry per failed ingestion event.
2. We develop a taxonomy of error categories—missing required or recommended fields, issuer ID mismatches relative to the CMS MR-PUF, invalid JSON, network/TLS failures, and schema anomalies—and quantify their prevalence across QHP API sources, years (with emphasis on 2025–2026), issuers, states, and endpoints.
3. Using the QHP specification [11], CMS Exchange PUF documentation [5, 6], and the Healthcare MRF API import model [8], we analyze why the fields that are most frequently missing (`formulary`, `cost_sharing`, identifiers, tiers, policy flags) are structurally necessary, and how their absence undermines not only QHP-level tools but also PUF- and MRF-based rate analytics.

## 2 Methods

### 2.1 Data Sources and Time Frame

Our primary empirical source is the HealthPorta Healthcare Data Dashboard, which exposes a roster of marketplace issuers at:

https://www.healthporta.com/healthcare-data/.

For each issuer, the dashboard publishes an import log page at `/healthcare-data/issuer/<slug>/import/`, summarizing ingestion attempts and listing individual error events in an “Error log” table. Each import reflects an attempt to ingest the issuer’s QHP Provider & Formulary API files—conceptually aligned with `index.json`, `plans.json`, `providers.json`, `drugs.json`, and closely related JSON resources defined in the QHP specification [11].

We captured a snapshot of all issuer import logs in early December 2025. At that time, the issuer roster comprised 735 entities. Of these, 339 issuers had at least one recorded error, yielding 59,899 error rows in total. These error rows represent failures encountered while processing QHP API artifacts: plan JSON files, provider directories, drug formularies, and index JSONs.

In addition to HealthPorta’s logs, we rely on CMS’s Exchange Public Use Files—specifically the Machine-readable URL PUF (MR-PUF)—to define the canonical universe of issuer-level index URLs and issuer identifiers for each plan year. CMS describes the MR-PUF as a file that “contains issuer-level data identifying machine-readable index URLs that lead to the issuer’s provider, formulary, and plan JSON URLs,” with one record per issuer reporting its index URL and a technical point of contact [6]. We focus on the 2025 and 2026 MR-PUFs and treat their State, Issuer ID, and URL Submitted fields as the authoritative mapping from issuer IDs to machine-readable index URLs for those years.

Although our main goal is to understand current coverage years, we did not filter older records out of the raw data. Instead, we record all years present in issuer files. As a result, our dataset also contains a modest number of errors associated with earlier plan years (2018–2024). In most cases, these older-year errors arise because issuers continue to host JSON records for prior coverage years alongside current-year data, or have not segregated historical content from current QHP API endpoints.

### 2.2 Scraping and Parsing

We programmatically fetched each issuer’s import page and parsed the HTML table under the “Error log” heading. Each error is represented in the page as a pair of rows:

- A header row containing: log identifier (`log_id`), error type (always `err` in this analysis), source (`plans`, `providers`, `drugs`, `json_index`, `formulary`), level (`json` or other), and file URL (`file_url`) pointing to the issuer-hosted QHP API resource; and
- A detail row containing the free-text error message.

We stored each error event as a JSON object with the following fields: issuer slug, issuer name, `log_id`, type, source, level, `file_url`, free-text message, and the canonical import log URL (`import_url`). These objects are aggregated into derived tables described below.

### 2.3 Error Taxonomy

To make the analysis tractable, we mapped free-text error messages into a controlled set of categories using pattern matching on the message text. The primary categories are:

- **Missing recommended field:** `cost_sharing` — messages indicating that the recommended `cost_sharing` field is missing or incorrect in a plan file.
- **Missing required field:** `formulary` — messages indicating that the required `formulary` field is missing or incorrect.
- **Missing required field (other/drug)** — other messages indicating missing required fields, including a specific subtype for missing required drug fields.
- **Issuer ID mismatch vs index** — messages indicating that a file describes an issuer that is not defined or allowed by the CMS machine-readable URL index for that plan year.
- **Invalid JSON in index or data file** — messages indicating JSON parsing failures, invalid characters, or incomplete JSON.
- **Network/SSL failure** — messages indicating network connectivity or TLS issues (e.g., certificate verification failures, handshake errors).
- **HTTP error from source** — messages indicating HTTP error responses while downloading a resource.
- **Plan ID length** *>* 14 — messages indicating that a plan identifier exceeds the expected length.
- **Schema violation / Other** — residual messages that do not fit the above categories.

We then constructed aggregate tables summarizing error counts by category, source, year, issuer, state, and endpoint. The global category distribution is stored in `error_categories.xls`.

### 2.4 JSON Error Subtypes

Within the “Invalid JSON in index or data file” category, we further refined errors into JSON subtypes, stored in a `json_detail` field and summarized in `json_subtype_by_source.xls` and `json_error_breakdown.xls`. These subtypes include:

- HTML or non-JSON returned — the endpoint returns HTML (often an error page) instead of JSON.
- Invalid or non-UTF-8 bytes — parsing fails due to invalid UTF-8 byte sequences.
- Truncated/incomplete JSON — messages explicitly indicating incomplete or truncated JSON.
- Lexical/parse errors — generic parsing errors that cannot be attributed to the above more specific patterns.

We additionally grouped identical JSON error messages and endpoints in `json_error_breakdown.xls` to reveal recurring patterns across issuers and URLs.

### 2.5 Derived Aggregations

To support the quantitative analysis reported in Section 3, we generated the following derived tables from the line-delimited error data (`all_errors.jsonl`):

- **Global error categories** (`error_categories.xls`) — counts of errors per category, including number of affected issuers and share of total errors.
- **Category** *×* **source** (`category_by_source.xls`) — counts of errors per category split across `plans`, `providers`, `drugs`, `json_index`, and `formulary` sources.
- **Category** *×* **year** (`category_by_year.xls`) — counts per category and plan year extracted from the error message text.
- **Issuer-level counts** (`issuers_with_errors.xls`, `top_issuers.xls`) — total errors per issuer and a ranked subset of issuers with the largest error counts.
- **State roll-up** (`state_rollup.xls`) — total errors per issuer state, derived from the issuer roster table.
- **JSON error summaries** (`json_error_breakdown.xls`, `json_subtype_by_source.xls`) — subtypes and sources of JSON errors.
- **Network and TLS failures** (`network_ssl_details.xls`) — per-issuer network/TLS failure details.
- **Plan ID anomalies** (`plan_id_anomalies.xls`) — rare cases of plan IDs exceeding the expected length.

Each of these spreadsheets includes one or more columns with URLs pointing back to HealthPorta issuer import logs and source file URLs, preserving a direct link from aggregate statistics to the original web-based evidence.

### 2.6 Healthcare MRF API and Import Pipeline

Our analysis of QHP Provider & Formulary API quality is tightly coupled to a separate but related system: the Healthcare MRF API [8]. This open-source Python service ingests CMS marketplace machine-readable files and issuer-hosted transparency data (including plan and rate files), normalizes them into a PostgreSQL database, and exposes the results through a REST API. The API is an example of the kind of specialized tooling that CMS envisions when it notes that health plan price transparency data are intended to be used by experienced third parties to help consumers interpret costs [10]. Rather than expecting individual patients to download large, complex machine-readable files, the pipeline turns them into plan-centric and provider-centric views that can be exposed through a consumer-facing dashboard. In a typical deployment:

- PostgreSQL and Redis instances are provisioned.
- The service is configured via a `.env` file based on `.env.example`.
- Imports are triggered by running a command such as `python main.py start mrf`, which enqueues MRF import work.
- A worker process (e.g., `python main.py worker process.MRF –burst`) downloads, parses, and loads the machine-readable files.
- The workflow is usually scheduled via cron (for example, weekly) to keep the MRF data aligned with CMS and issuer updates.

After the first import, MRF data become available on the API port (default 8080), and the import status can be monitored via endpoints such as:

- `/api/v1/import` — high-level status of current and past imports.
- `/api/v1/plan/id/{PLAN_ID}` — plan-centric views combining MRF data for a specific marketplace plan.

In production, HealthPorta uses this MRF pipeline to power its ACA Healthcare Data Dashboard [12], which tracks imports from CMS marketplace MRFs via the Healthcare MRF API and reports plan counts, issuer counts, and import errors per issuer. HealthPorta’s public plan profiles combine official CMS filings, MRF rates, plan metadata, formulary tiers, deductible and MOOP summaries, and issuer-level drug and provider insights.

### 2.7 Schema Context: Role of Key QHP Fields

To interpret the impact of observed errors, we rely on the QHP Provider & Formulary API specification [11], which defines the role of several key fields:

- In `plans.json`:
  - `plan_id_type`, `plan_id`, and issuer IDs serve as primary keys across files.
  - `network` lists network tiers; each network includes `network_tier` values.
  - `formulary` lists formularies associated with the plan; each formulary includes `drug_tier`, `mail_order`, and optionally `cost_sharing`.
- Within a `formulary` entry:
  - `drug_tier` (required) names the tier (e.g., GENERIC, PREFERRED, SPECIALTY).
  - `mail_order` (required) indicates whether mail-order is available for this tier.
  - `cost_sharing` (optional array) holds cost-sharing rules for that tier.
- Within each `cost_sharing` element:
  - `pharmacy_type`, `copay_amount`, `copay_opt`, `coinsurance_rate`, and `coinsurance_opt` are required if `cost_sharing` is present (for example, 1-MONTH-IN-RETAIL with a fixed copay before deductible). In the context of health plan price transparency and TiC MRF data, these fields are the bridge between negotiated rates and member-facing estimates of out-of-pocket spending for specific pharmacy channels [13, 14].
- In `providers.json`:
  - Each provider entry includes a `plans` array referencing `plan_id` and `network_tier` values that must match those declared in `plans.json`.
- In `drugs.json`:
  - Required: `rxnorm_id`, `drug_name`, `plan_id`, `drug_tier`.
  - Optional but important: `prior_authorization`, `step_therapy`, `quantity_limit` flags that indicate utilization-management policies.

We will repeatedly refer back to this schema context when interpreting the missing fields we observe—especially the absence of `formulary` and `cost_sharing`, mismatched IDs, and missing tiers and flags.

## 3 Results

### 3.1 Overall Error Prevalence

Among 735 issuers listed in the HealthPorta dashboard, 339 (46.1%) have at least one recorded import error in our December 2025 snapshot. Across all issuers and sources, we observe 59,899 error events in total, as summarized in the aggregate table `error_categories.xls`.

The global distribution of categories is highly skewed. Three categories dominate:

- missing recommended `cost_sharing` fields;
- missing required `formulary` fields; and
- issuer ID mismatches relative to the CMS machine-readable URL index.

**Figure 1:**
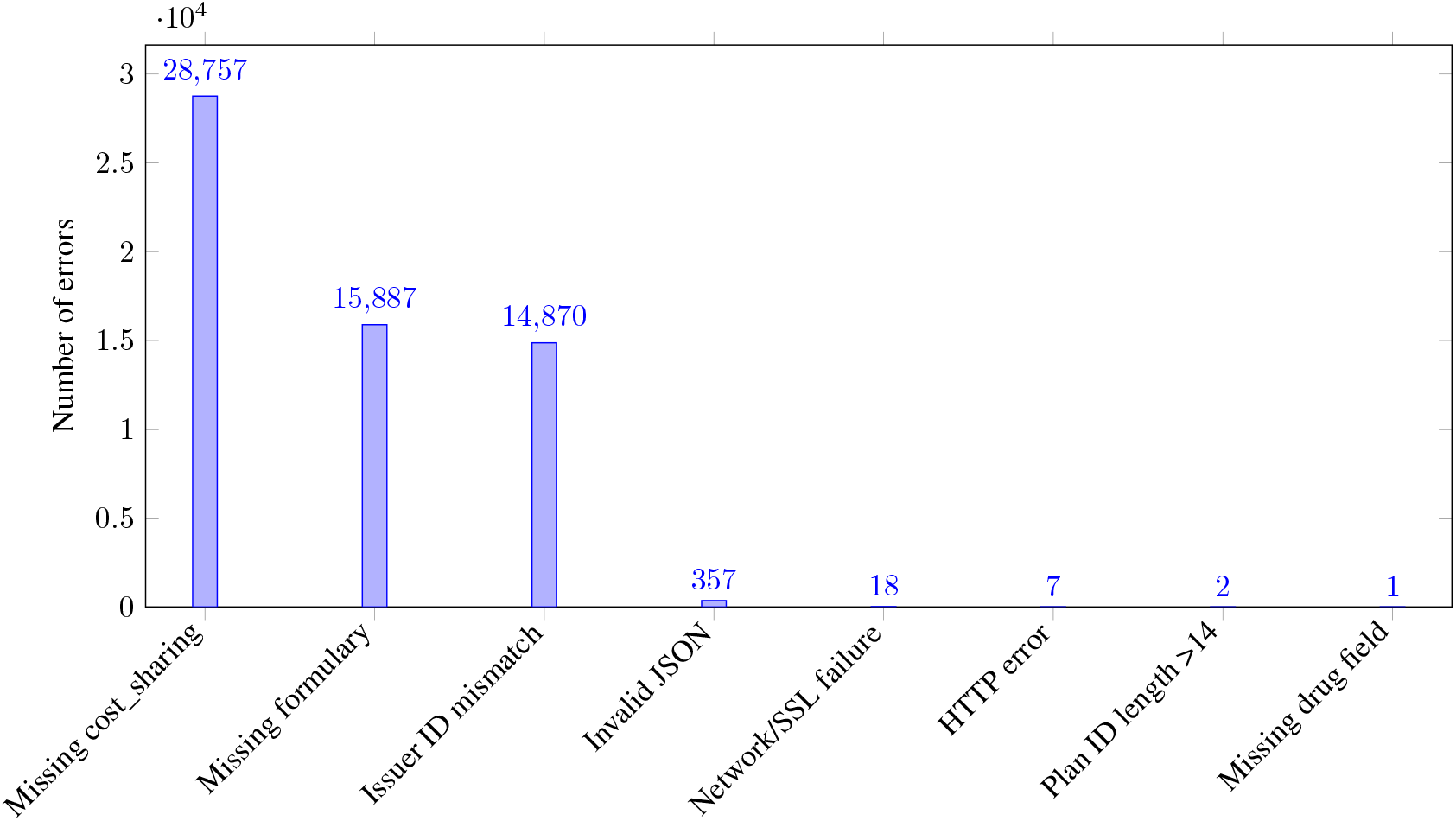
Global distribution of error categories across all 59,899 issuer-side QHP API errors.

In the broader price-transparency literature, similar patterns emerge: analyses of hospital and insurer transparency files under the hospital price transparency and Transparency in Coverage rules emphasize that the main obstacles are not simply file availability but file size, structural complexity, and lack of standardization and completeness in key fields [14, 15, 13]. Our findings extend this picture to marketplace QHP provider and formulary APIs, where missing core fields and identifier mismatches, rather than transport errors, account for nearly all observed failures.

**Table 1:** Global error categories (from error_categories.xls).

| Category | Errors | Issuers | Share |
| --- | --- | --- | --- |
| Missing recommended field: `cost_sharing` | 28757 | 148 | 48.01% |
| Missing required field: `formulary` | 15887 | 69 | 26.52% |
| Issuer ID mismatch vs index | 14870 | 179 | 24.83% |
| Invalid JSON in index or data file | 357 | 87 | 0.60% |
| Network/SSL failure | 18 | 18 | 0.03% |
| HTTP error from source | 7 | 4 | 0.01% |
| Plan ID length > 14 | 2 | 1 | 0.00% |
| Missing required field (drug) | 1 | 1 | 0.00% |

### 3.2 Error Categories by Source

To understand where issuer pipelines are failing, we break errors down by file source (`plans`, `providers`, `drugs`, `json_index`, `formulary`) using `category_by_source.xls`. As expected, most structural schema violations are concentrated in plan files:

- all 28,757 missing `cost_sharing` errors occur in plan files;
- all 15,887 missing `formulary` errors occur in plan files;
- all 14,870 issuer ID mismatch errors arise from plan files.

By contrast, JSON-formatting and transport-related problems are more prominent in provider and index files. Invalid JSON is distributed as 268 errors from provider files, 45 from plan files, and 44 from index files, with network/TLS and HTTP errors mostly affecting index and provider endpoints.

**Table 2:** Error categories by source (from category_by_source.xls).

| Category | Source | Errors |
| --- | --- | --- |
| Missing recommended field: `cost_sharing` | plans | 28757 |
| Missing required field: `formulary` | plans | 15887 |
| Issuer ID mismatch vs index | plans | 14870 |
| Invalid JSON in index or data file | providers | 268 |
| Invalid JSON in index or data file | plans | 45 |
| Invalid JSON in index or data file | json_index | 44 |
| Network/SSL failure | json_index | 18 |
| HTTP error from source | providers | 6 |
| Plan ID length > 14 | plans | 2 |
| HTTP error from source | formulary | 1 |
| Missing required field (drug) | formulary | 1 |

### 3.3 Error Categories by Year

Using the `category_by_year.xls` table, we examine how error categories evolve across plan years. Because our main goal is to understand current coverage years, we focus on 2025 and 2026, but explicitly report the smaller tail of errors associated with older plan years.

For the dominant categories, we observe:

- Missing `cost_sharing`:
  - **–** 2018–2023 combined: 931 errors (11 + 9 + 8 + 9 + 296 + 598).
  - **–** 2024: 1,574 errors.
  - **–** 2025: 13,583 errors.
  - **–** 2026: 12,669 errors.
- Missing `formulary`:
  - **–** 2024: 23 errors.
  - **–** 2025: 7,630 errors.
  - **–** 2026: 8,234 errors.
- Issuer ID mismatch vs index:
  - **–** 2022–2024 combined: 1,523 errors (698 + 401 + 424).
  - **–** 2025: 6,978 errors.
  - **–** 2026: 6,369 errors.

**Table 3:** Error categories by plan year (from category_by_year.xls).

| Category | Year | Errors |
| --- | --- | --- |
| Missing recommended field: `cost_sharing` | 2018 | 11 |
| Missing recommended field: `cost_sharing` | 2019 | 9 |
| Missing recommended field: `cost_sharing` | 2020 | 8 |
| Missing recommended field: `cost_sharing` | 2021 | 9 |
| Missing recommended field: `cost_sharing` | 2022 | 296 |
| Missing recommended field: `cost_sharing` | 2023 | 598 |
| Missing recommended field: `cost_sharing` | 2024 | 1574 |
| Missing recommended field: `cost_sharing` | 2025 | 13583 |
| Missing recommended field: `cost_sharing` | 2026 | 12669 |
| Missing required field: `formulary` | 2024 | 23 |
| Missing required field: `formulary` | 2025 | 7630 |
| Missing required field: `formulary` | 2026 | 8234 |
| Issuer ID mismatch vs index | 2022 | 698 |
| Issuer ID mismatch vs index | 2023 | 401 |
| Issuer ID mismatch vs index | 2024 | 424 |
| Issuer ID mismatch vs index | 2025 | 6978 |
| Issuer ID mismatch vs index | 2026 | 6369 |
| Plan ID length > 14 | 2025 | 1 |
| Plan ID length > 14 | 2026 | 1 |

Aggregating across all categories:

- 2025 and 2026 together account for the vast majority of errors (28,192 and 27,273 errors respectively), reflecting current and upcoming coverage years.
- Earlier years (2018–2024) contribute a long but relatively small tail of errors. These arise because some issuers still expose multi-year or historical plan records in their QHP API JSON files instead of limiting the content strictly to the current coverage year.

### 3.4 Issuer-Level Patterns

Issuer-level views from `issuers_with_errors.xls` and `top_issuers.xls` reveal a heavy-tailed distribution of errors across issuers. Among the 735 issuers in the roster, 339 have at least one recorded error.

Across these 339 issuers:

- the median issuer has 94 errors;
- the 25th and 75th percentiles are 14 and 293.5 errors, respectively;
- the 95th percentile issuer has 577 errors; and
- the maximum is 1,092 errors for a single issuer.

### 3.5 Geographic Distribution

By joining issuers to their primary state in the HealthPorta roster and aggregating error counts by state (`state_rollup.xls`), we obtain a coarse geographic distribution of data-quality problems. Not surprisingly, states hosting large national carriers accumulate more errors, but there is substantial variation even after accounting for issuer size.

**Table 4:** Top issuers by error count (from top_issuers.xls).

| Rank | Issuer | Errors |
| --- | --- | --- |
| 1 | Blue Cross Blue Shield of Texas | 1092 |
| 2 | Anthem Health Plans of New Hampshire | 803 |
| 3 | Anthem Insurance Companies, Inc. | 803 |
| 4 | Blue Cross Blue Shield of Wisconsin (PPO and out of network POS) | 803 |
| 5 | Community Insurance Company | 803 |
| 6 | Compcare Health Services Insurance Corporation (HMO/POS in network) | 803 |
| 7 | Healthy Alliance Life Insurance Company | 803 |
| 8 | Matthew Thornton Health Plan, Inc. | 803 |
| 9 | Simply Healthcare Plans, Inc. doing business as Wellpoint Florida, Inc. | 803 |
| 10 | Wellpoint Insurance Company | 803 |

**Table 5:** Top states by issuer error count (from state_rollup.xls).

| State | Errors |
| --- | --- |
| TX | 5385 |
| WI | 3935 |
| FL | 3821 |
| OH | 3289 |
| IN | 2859 |
| MO | 2749 |
| NH | 2352 |
| MI | 2345 |
| AZ | 2228 |
| UT | 2212 |

**Table 6:** JSON error subtypes (from json_error_breakdown.xls).

| JSON error subtype | Errors | Issuers |
| --- | --- | --- |
| Invalid UTF-8 bytes in JSON | 230 | 20 |
| HTML or non-JSON returned | 124 | 64 |
| Truncated/incomplete JSON | 3 | 3 |

### 3.6 Endpoint-Level Failures

Endpoint-level statistics, derived by grouping errors by `file_url`, show that a relatively small set of URLs are responsible for large clusters of failures. Some plan JSON endpoints accumulate hundreds or thousands of missing-field or issuer ID mismatch errors, indicating that the underlying data are systematically mis-specified rather than intermittently failing.

Provider and index URLs are overrepresented among endpoints associated with invalid JSON or HTML responses, suggesting that some QHP API endpoints are implemented as thin wrappers around internal web applications, error pages, or authentication layers rather than as stable JSON feeds.

These endpoint-level views are directly actionable for remediation: they pinpoint specific URLs that issuers or vendors can test and fix to eliminate large pockets of ingestion failures.

### 3.7 JSON Errors in Detail

JSON-related failures are numerically small (357 events in total) but provide insight into the robustness of issuer-hosted APIs and file servers. Using `json_error_breakdown.xls`, we classify these errors into three subtypes:

- invalid UTF-8 bytes in JSON — 230 errors across 20 issuers;
- HTML or non-JSON returned — 124 errors across 64 issuers;
- truncated or incomplete JSON — 3 errors across 3 issuers.

The `json_subtype_by_source.xls` table further decomposes these JSON error subtypes by file source (Table 7). Invalid UTF-8 bytes are almost entirely confined to provider files, whereas HTML responses are spread across index, provider, and plan endpoints, and truncation errors appear only in plan files.

**Table 7:** JSON error subtypes by source (from json_subtype_by_source.xls).

| JSON error subtype | Source | Errors | Issuers |
| --- | --- | --- | --- |
| Invalid UTF-8 bytes in JSON | providers | 228 | 18 |
| Invalid UTF-8 bytes in JSON | plans | 2 | 2 |
| HTML or non-JSON returned | json_index | 44 | 44 |
| HTML or non-JSON returned | providers | 40 | 20 |
| HTML or non-JSON returned | plans | 40 | 20 |
| Truncated/incomplete JSON | plans | 3 | 3 |

**Table 8:** Network and TLS failures (from network_ssl_details.xls).

| Issuer | Source | Message (abridged) |
| --- | --- | --- |
| Blue Cross Blue Shield of Arizona, Inc. | json_index | certificate verify failed (e.g., self-signed or untrusted CA) |
| BlueCross BlueShield of Tennessee, Inc. | json_index | certificate verify failed or handshake error at JSON index endpoint |
| Dominion National | json_index | repeated handshake/certificate errors across multiple index URLs |
| Health Alliance Medical Plans, Inc. | json_index | TLS handshake failure when fetching JSON index |
| HMO Louisiana, Inc. | json_index | certificate verification error when connecting to index URL |
| InStil Health Insurance Company | json_index | TLS error retrieving index file |
| Louisiana Health Service & Indemnity Co. | json_index | certificate or protocol error at index endpoint |
| Paramount Insurance Company | json_index | timeout or unreachable host for index URL |
| Summa Insurance Company | json_index | timeout or unreachable host for index URL |

**Table 9:** Plan ID length anomalies (from plan_id_anomalies.xls).

| Issuer | Plan ID | Year |
| --- | --- | --- |
| Select Health of South Carolina, Inc. | 73107SC00100010 | 2025 |
| Select Health of South Carolina, Inc. | 73107SC00100010 | 2026 |

These issues echo prior evaluations of price transparency data, which report that compliant machine-readable files often require substantial cleaning to handle encoding problems, schema differences, and partial responses before they can support downstream analytics or consumer tools [14, 15].

### 3.8 Network and TLS Failures

The `network_ssl_details.xls` table lists 18 network-or TLS-related failures across 9 issuers. Typical messages include certificate verification failures (self-signed or mis-chained certificates), handshake errors, and connection timeouts.

These issues are concentrated at JSON index endpoints. For example, some issuers experience repeated failures when connecting to their QHP index URLs, and others account for multiple TLS failures across distinct issuer identifiers.

### 3.9 Plan ID and Schema Anomalies

Finally, `plan_id_anomalies.xls` captures rare schema anomalies in plan identifiers. We observe two instances where a plan ID exceeds the expected 14-character limit:

- both anomalies involve the same issuer, Select Health of South Carolina, Inc.;
- the offending plan ID is `73107SC00100010`, appearing in both 2025 and 2026 plan files.

## 4 Discussion

### 4.1 Schema Context and the Necessity of Key Fields

The QHP schema is designed so that a small set of fields act as load-bearing connectors across files:

- `plan_id` and issuer IDs link `plans.json`, `providers.json`, `drugs.json`, and CMS indices or PUFs.
- `formulary` in `plans.json` defines the list of formularies and tiers associated with each plan.
- Within each `formulary`, `drug_tier` and `mail_order` define tier semantics and mail-order availability.
- `cost_sharing` within the formulary defines, for each `pharmacy_type`, the copay and/or coinsurance structure and its relationship to the deductible.
- In `providers.json`, `plan_id` and `network_tier` link providers to specific plan tiers.
- In `drugs.json`, `plan_id`, `drug_tier`, and the optional flags `prior_authorization`, `step_therapy`, and `quantity_limit` encode coverage and utilization management at the drug level.

We highlight three especially crucial pieces:

1. `formulary` (required in `plans.json`) — the per-plan menu of drug tiers. Without it, `drugs.json` still lists `drug_tier` values, but they float in isolation, with no canonical list of tiers per plan or tier-level summary.
2. `cost_sharing` (optional in schema, functionally mandatory) — the bridge from tier labels to actual dollars and percentages. Without it, tiers become nominal labels with no numeric meaning, and cost comparisons are impossible.
3. Identifiers and foreign keys (`plan_id`, issuer IDs, `network_tier`, `drug_tier`) — the keys that tie plans, providers, drugs, PUFs, and MRFs together. When they are missing or mismatched, joins fail and analyses become incomplete or misassigned.

Optional but crucial flags (`prior_authorization`, `step_therapy`, `quantity_limit`) operate similarly: their absence creates ambiguity—does the plan truly have no restrictions, or did the issuer simply omit the field?

### 4.2 Dominant Issuer-Side Problems

Against this schema backdrop, our empirical results show that three issues dominate the error landscape:

- Omitted `cost_sharing`: 28,757 errors (48.0% of all observed errors).
- Omitted `formulary`: 15,887 errors (26.5%).
- Issuer ID mismatches: 14,870 errors (24.8%) relative to the CMS machine-readable URL index.

These are not marginal aspects of the specification:

- Missing `formulary` removes the plan-level summary and validation surface for drug tiers.
- Missing `cost_sharing` removes the numeric meaning of tiers and undermines cost comparisons.
- ID mismatches prevent safe joins across QHP files, CMS Exchange PUFs, and MRF data.

The fact that nearly all observed errors fall into these three categories suggests that many issuers treat these fields as optional or informational rather than structurally necessary, and/or lack rigorous validation at data production time.

### 4.3 Structural Versus Operational Failures

Our taxonomy distinguishes between structural schema violations and operational failures in QHP APIs:

- **Structural issues** (missing fields, ID mismatches, plan ID anomalies) are persistent properties of the underlying data. They typically affect many plans for a given issuer and persist across import attempts and across years— especially in the 2025–2026 coverage years that dominate our dataset.
- **Operational issues** (invalid JSON, HTML responses, network/TLS errors) often reflect configuration or deployment problems. While they may also persist over time, they are conceptually simpler to detect and fix with proper monitoring and deployment practices.

The dominance of structural issues implies that simply improving uptime or HTTP status codes is not enough. Issuers and vendors must invest in schema-aware validation and cross-file consistency checks, not just endpoint availability.

### 4.4 Implications for QHP-Only Consumers

For tools and analyses that use QHP provider and formulary APIs in isolation:

- Missing `formulary` means that drug-level coverage appears only as `drug_tier` tags in `drugs.json`, with no canonical tier set or tier-level descriptions. This blocks tier-level analytics.
- Missing `cost_sharing` means there is no machine-readable bridge from tier labels to out-of-pocket amounts. Consumer-facing tools must fall back to non-structured documents.
- Missing or ambiguous utilization-management flags (`prior_authorization`, `step_therapy`, `quantity_limit`) undermine fairness and access analyses.
- ID and tier mismatches prevent reliable mapping of providers to plans and tiers, and of drugs to plans and tiers, weakening network adequacy metrics and drug-coverage comparisons.

In short, even without considering MRF data, the missing QHP fields we observe render many of the specification’s intended use cases (tier comparisons, access analysis, decision support) impossible or unreliable. These concerns are consistent with prior evidence that inaccurate provider directories are associated with worse access and a higher risk of surprise out-of-network bills [3, 2]. More broadly, they mirror findings from the price-transparency literature that the mere existence of machine-readable price data is insufficient: patients only benefit when data are complete enough for tools to surface meaningful, plan-specific cost information [16, 13].

### 4.5 Interaction with CMS Exchange PUFs and the Healthcare MRF API

The QHP Provider & Formulary APIs, CMS Exchange PUFs, and the Healthcare MRF API form a composite data pipeline:

- QHP APIs define who is covered and under what benefit structure:
  - `plans.json` *→* plan identifiers, marketing names, network tiers, formularies, and recommended `cost_sharing` structures.
  - `providers.json` *→* which providers participate in which plan networks and tiers.
  - `drugs.json` *→* which drugs are covered on which tiers, plus utilization-management flags.
- Exchange PUFs provide plan- and issuer-level metadata across multiple datasets (Benefits and Cost Sharing, Rate, Plan Attributes, Service Area, Network, Plan ID Crosswalk, Machine-readable URL, Transparency in Coverage, and others) for plan years 2014–2026 [5, 6].
- The Healthcare MRF API ingests rate transparency MRFs from CMS and issuers, and ties negotiated rates and allowed amounts back to plans and issuers defined by PUFs and QHP APIs [8].

Because these systems depend on shared identifiers and shared semantics, the missing QHP fields we observe have direct consequences for PUF- and MRF-based analytics:

1. **Missing** `formulary` **blocks break plan-level drug views in the PUF/MRF layer**. The Healthcare MRF API can store and serve negotiated rates for prescription drugs associated with a plan, and plan-level metadata can be enriched with PUF information (e.g., rates, service areas, quality). But without `formulary` in `plans.json` there is no structured map of which tiers exist for that plan or how those tiers behave. Plan-level drug coverage views devolve into lists of prices without coherent tier context.
2. **Missing** `cost_sharing` **makes MRF prices hard to translate into member out-of-pocket costs**. MRFs generally describe allowed amounts and negotiated rates between issuers and providers or pharmacies. To answer member-facing questions (“What will I pay at the pharmacy?”), a tool must combine MRF negotiated-rate data with QHP `cost_sharing` rules at the relevant tier and pharmacy channel. When `cost_sharing` is absent, applications cannot convert negotiated rates into member out-of-pocket estimates.
3. **Issuer ID and** `plan_id` **mismatches break joins between QHP files, Exchange PUFs, and MRF datasets**. The MR-PUF organizes issuer-level machine-readable index URLs by state and five-digit HIOS issuer ID, and CMS also publishes Plan ID Crosswalk and other PUFs that attach plan-level metadata to plan identifiers [5, 6]. When QHP plan files contain issuer IDs or plan IDs that do not match these CMS indices, some plans present in MRF data cannot be matched to QHP or PUF metadata (or vice versa), providers and drug coverage cannot be consistently aligned with pricing, and issuer- and state-level statistics based on joined QHP+PUF+MRF data become biased or incomplete.
4. **Legacy plan years in QHP files complicate PUF/MRF imports and plan history**. Exchange PUFs and MRFs are organized by plan year. When QHP files intermix 2018–2024 plans with 2025–2026 plans at the same endpoints, it becomes difficult to ensure that MRF records for a given `plan_id` and year correspond to the correct benefits, networks, and formularies.
5. **Provider and network tier issues weaken MRF-based network adequacy views**. Network adequacy and access analyses require mapping providers to networks and networks to plans. If QHP provider or network data have missing or inconsistent `network_tier` or `plan_id` values, any composite metric that combines cost (from MRFs) and access (from QHP networks and PUFs) becomes fragile.

Taken together, the same small set of missing QHP fields—`formulary`, `cost_sharing`, and correctly keyed identifiers and tiers—are structural prerequisites for Exchange PUFs and the Healthcare MRF API to produce joined QHP+PUF+MRF views at scale. When they are absent or inconsistent, the MRF importer can still ingest rates, and PUFs still provide high-level metadata, but their outputs cannot be safely attached to plan benefits, provider networks, or formulary tiers, blunting the impact of otherwise rich transparency datasets. These findings sit squarely within a broader policy conversation about how to build more reliable, national-scale provider directory infrastructure [17]. They also complement early empirical work showing that when insurer and hospital transparency datasets can be reliably linked, negotiated prices across sources tend to be reasonably concordant, suggesting that data quality and cross-dataset connectivity, rather than fundamental unreliability of prices, are the main bottlenecks [18, 14].

Our issuer-level error counts should therefore be interpreted as indicators of observed data-quality issues in a single importer pipeline, not as formal regulatory findings. They are intended to support remediation and improved transparency, rather than to attribute fault or non-compliance.

### 4.6 Open transparency datasets and clarity of US health insurance

Federal price-transparency initiatives are motivated by the goal of giving patients, employers, and other purchasers clearer information about what care will cost. CMS describes health plan price transparency as a way to help consumers know the cost of a covered item or service before receiving care and explicitly notes that the machine-readable files required under the Transparency in Coverage rule are intended to be used by third parties, such as researchers and application developers, to help consumers interpret prices [9, 10]. Bernstein and Crowe’s narrative policy review of US price transparency efforts similarly argues that open pricing data are a necessary but not sufficient condition for empowering patients; usable tools and standardization are also required [13].

A growing empirical literature evaluates whether such transparency actually changes behavior or spending. Parente estimates that fully implemented price-transparency policies could reduce total spending for the privately insured US population by roughly 7%, corresponding to tens of billions of dollars in potential annual savings, provided that robust tools exist to convert raw prices into usable information for shoppable services [19]. Lin et al.’s systematic review of 87 US price-transparency studies finds mixed but generally promising evidence: when tools succeed in making prices salient and comparable at the point of decision, they can reduce prices and out-of-pocket spending for select services, although uptake remains limited and effects are heterogeneous [16].

At the same time, multiple studies emphasize that the current generation of machine-readable transparency files is often difficult to use in practice. Maleki et al. report that hospital transparency MRFs are extremely large and technically complex, requiring substantial preprocessing to extract usable prices, while a recent Congressional Research Service report catalogs technical challenges with private health insurance transparency data, including file size, format variation, and access barriers [14, 15]. Henderson and Mouslim show that insurer and hospital transparency datasets can provide mutually validating price information when they overlap, but that the overlap itself is surprisingly limited, further underscoring the importance of data completeness and alignment [18].

Our results add a complementary perspective from the QHP Provider & Formulary API ecosystem. Even when insurers publish MRFs that comply with federal rules, missing QHP fields such as `formulary`, `cost_sharing`, and clean plan identifiers can prevent transparency tools from translating negotiated rates into clear, plan-specific coverage stories for people. In this sense, issuer-side QHP data quality is a key mediating layer between the legal availability of MRF data and the practical clarity of health insurance for patients and purchasers.

### 4.7 Limitations and Future Work

This study analyzes a single cross-sectional snapshot of issuer import logs for QHP provider and formulary APIs, with a primary emphasis on the coverage years 2025 and 2026. Error patterns may evolve over time as issuers update their systems or as CMS requirements and schemas change. Our categorization also depends on HealthPorta’s ingestion processes and error messages, which may not capture all underlying problems and may themselves contain biases.

The presence of older plan years (2018–2024) in many issuers’ JSON files introduces additional complexity: it is often unclear whether these historical records are intentionally maintained for archival purposes or are simply remnants of past coverage years that were never removed. Future work could extend this approach by:

- analyzing error trends over time, separating current-year errors from legacy-year errors;
- comparing results with alternative QHP API validators or schema definitions;
- incorporating more Exchange PUF datasets (Benefits and Cost Sharing, Rate, Network, Plan ID Crosswalk) to understand how QHP API quality interacts with plan and issuer-level metadata; and
- conducting issuer- and vendor-level case studies to understand organizational and technical drivers of data quality.

## Data Availability

All data produced are available online at https://www.healthporta.com/healthcare-data/ and https://github.com/EndurantDevs/healthcare-mrf-api/raw/refs/heads/main/reports/errors-article-12-2025.zip

https://github.com/EndurantDevs/healthcare-mrf-api/raw/refs/heads/main/reports/errors-article-12-2025.zip

## Ethics

This study analyzes error logs and machine-readable files derived from public web resources (CMS Health Insurance Exchange Public Use Files, issuer-hosted Qualified Health Plan Provider & Formulary APIs, and transparency machine-readable files). No individual-level clinical, claims, or personally identifiable information was used. The analysis does not involve human subjects research as defined by U.S. regulations and did not require institutional review board approval.

## Funding

This work did not receive specific funding from any agency in the public, commercial, or not-for-profit sectors. The HealthPorta Healthcare Data Dashboard and Healthcare MRF API are maintained as open-source projects by EndurantDevs LLC.

## Competing interests

Dmytro Nikolayev is the founder of EndurantDevs LLC and maintains the HealthPorta Healthcare Data Dashboard and the Healthcare MRF API, which are used as data sources and infrastructure in this analysis. The author declares no other competing interests.

## 5 Data and Supplementary Materials

All aggregate statistics reported in this article are derived from the line-delimited error dataset (`all_errors.jsonl`) and a set of `.xls` workbooks included as supplementary materials:

- `error_categories.xls` — global error categories (counts, issuers, share of total errors, and an `Example_Log_URL` for each category).
- `category_by_source.xls` — error categories broken down by file source (`plans`, `providers`, `drugs`, `json_index`, `formulary`), with example log URLs.
- `category_by_year.xls` — error categories broken down by plan year (2018–2026), with example log URLs.
- `issuers_with_errors.xls` — per-issuer error counts and summary statistics, including `Import_Log_URL` for each issuer.
- `top_issuers.xls` — issuers with the largest error counts.
- `state_rollup.xls` — error counts aggregated by issuer state.
- `json_error_breakdown.xls` — overall JSON error subtypes and counts.
- `json_subtype_by_source.xls` — JSON error subtypes broken down by file source.
- `network_ssl_details.xls` — detailed network and TLS failures per issuer.
- `plan_id_anomalies.xls` — plan ID length anomalies and associated plan years.

Each of these spreadsheets includes one or more columns with URLs pointing back to a concrete HealthPorta error log or issuer-hosted QHP JSON file, enabling independent inspection and replication of the patterns described in the main text.

The full set of raw and derived files (`all_errors.jsonl` and the accompanying `.xls` workbooks) is distributed as a single archive, `errors-article-12-2025.zip`, in the Healthcare MRF API GitHub repository [20]. In particular, all data and analysis workbooks used in this article are openly available at:

## 6 Conclusion

Using issuer import logs from the HealthPorta Healthcare Data Dashboard, we analyzed 59,899 QHP provider and formulary API ingestion errors across 735 issuers, 339 of which had at least one failure. A narrow set of issuer-side problems—missing `cost_sharing` and `formulary` fields and issuer ID mismatches relative to the CMS Machine-readable URL PUF—accounts for almost all observed errors. JSON formatting issues, network/TLS failures, and rare schema anomalies provide additional evidence of operational fragility.

Our analysis shows that plan years 2025 and 2026 dominate both the volume of data and the volume of errors, but also that many issuers continue to expose legacy plan years in the same JSON files, creating a long tail of historical errors. Drawing on the QHP specification, CMS Exchange PUF documentation, and the Healthcare MRF API import model, we show that the fields most frequently missing—`formulary`, `cost_sharing`, identifiers, tiers, and policy flags—are structurally necessary for both QHP-only tools and for composite QHP+PUF+MRF analytics.

Substantial work remains for issuers and their vendors to make the QHP Provider & Formulary APIs, Exchange PUFs, and associated MRFs reliably usable in practice. Automated validation of required fields and identifiers, robust JSON and encoding checks, clearer handling of plan-year scope, and routine monitoring of endpoints and certificates would address the majority of observed problems and align with broader proposals for building more reliable national provider directory infrastructure [17]. Until such measures are widely adopted, researchers and regulators should treat QHP API–, PUF-, and MRF-derived insights with caution and explicitly account for non-trivial and uneven data loss across issuers, states, plan years, and file types.

Viewed against broader evidence on price transparency, our findings suggest that relatively targeted fixes—ensuring that QHP plan files consistently include `formulary` and `cost_sharing`, maintaining clean issuer and plan identifiers across QHP, PUF and MRF datasets, and enforcing basic JSON and TLS hygiene—would unlock much more of the policy value of existing transparency rules. If those elements were in place, open MRF and QHP data could support routine, plan-specific estimates of expected out-of-pocket costs, more accurate comparisons of network breadth and drug coverage, and automated regulatory audits of adequacy and parity, bringing the US system closer to the kind of usable price information envisioned by recent federal rulemaking and empirical studies of price transparency tools [9, 19, 16, 13]. In that sense, improving issuer-side QHP data quality is not just an implementation detail; it is a necessary precondition for the promise of open machine-readable rate and benefit data to translate into clearer, more transparent insurance for people in the United States.

## Notes

### Competing Interest Statement

The authors have declared no competing interest.

### Funding Statement

This study did not receive any funding.
This work did not receive specific funding from any agency in the public, commercial, or not-for-profit sectors.
The HealthPorta Healthcare Data Dashboard and Healthcare MRF API are maintained as open-source projects by EndurantDevs LLC.

